# Spectral Validity and Spindle Detection of Wearable Frontal EEG: A Per-Subject Calibration Framework and Systematic Validation Against Polysomnography Using the Wearanize+ Dataset

**DOI:** 10.64898/2026.06.01.26354593

**Authors:** Yildiz Dilara Parry, Giovanni Briganti

**Author notes:** Corresponding author: Yildiz Dilara Parry.

## Abstract

Wearable EEG devices enable home sleep monitoring but require systematic spectral validation before their physiological outputs can serve as proxies for polysomnographic features. This study provides comprehensive spectral validation of the Zmax EEG headband against concurrent PSG using the Wearanize+ dataset. Seventy-one participants with adequate signal quality underwent simultaneous home PSG and Zmax recording. Bandpower correspondence, calibration robustness, within-subject reliability, lateralisation, and spindle detection were evaluated across all sleep stages. Absolute delta power values reported in this study were computed using 4-second FFT windows and should not be directly compared to normative values derived from 8-second windows, as the shorter window introduces a systematic positive offset in the delta band that does not cancel in absolute comparisons. Zmax systematically underestimates bandpower across all frequency bands (bias –0.41 to –0.74 log units), attributable to the active Fpz reference electrode. A per-subject N2-referenced calibration eliminates this bias; N2 calibration outperformed N3 and REM alternatives (mean post-calibration r=0.601 vs 0.479 and 0.489). Post-calibration spectral correspondence was strong for alpha (N3: r=0.806) and sigma (N3: r=0.752). Within-subject reliability was excellent (split-half r>0.99). Demographic factors explained less than 4% of offset variance. Lateralisation analysis was underpowered (36–39% power; N=194 required for 80% power). Spindle under-detection was traced to YASA’s relative sigma power pre-filter; lowering this threshold recovered PSG-equivalent counts with near-zero bias. These findings establish a validated calibration framework and evidence-based feature selection recommendations for Zmax-based sleep biomarker research.

## 1. Introduction

The quantification of sleep EEG features; spectral power in frequency bands associated with specific sleep stages, sleep spindle density, and slow oscillation amplitude provides clinically and scientifically meaningful information about neural, metabolic, and psychiatric health (Li et al., 2016; Lustenberger et al., 2012; Medina et al., 2014; Plante, 2021; Stango et al., 2025; The 23andMe Research Team et al., 2019; Wamsley et al., 2012). These features serve as candidate biomarkers for a range of conditions including depression, anxiety, insomnia, and neurodegenerative disease, and have emerged as primary targets for personalised medicine applications in sleep research. However, their measurement has historically depended on laboratory polysomnography (PSG), whose resource intensity and ecological constraints limit their utility for population-scale studies (Chinoy et al., 2021).

Consumer wearable EEG devices, including the Zmax headband, offer a scalable alternative. The Zmax records bilateral frontal EEG from F7-Fpz and F8-Fpz bipolar derivations at 256 Hz in a home-use format, and has been validated for automated sleep staging in laboratory settings (Esfahani et al., 2023). However, spectral validation as the systematic comparison of EEG bandpower and oscillatory features between Zmax and PSG has not been performed at population scale in home-recording conditions. Two fundamental questions remain unanswered: (1) does the Zmax preserve inter-individual variation in sleep EEG spectral features well enough to serve as a calibrated proxy for PSG-grade measurements, and (2) are there systematic biases that can be characterised and corrected?

The Wearanize+ dataset (Sikder et al., 2026) provides the empirical conditions to address both questions, comprising synchronised full PSG and Zmax recordings from 130 home-based participants, 100 of whom are available in the PlugNPlay release. This study performs systematic spectral validation using 71 subjects with adequate signal quality, examining bandpower correspondence across five frequency bands and all sleep stages, developing a per-subject calibration protocol, validating spindle density detection, and testing the feasibility of bilateral asymmetry analyses. Together these findings provide a complete methodological foundation for wearable-based sleep biomarker research.

## 2. Methods

### 2.1 Dataset and Participants

Wearanize+ PlugNPlay dataset is used for this study. From the 74 participants classified as having adequate signal quality (OK group) in the companion signal quality analysis, 71 had the required PSG frontal channels (F3:A2, F4:A1, C3:A2, C4:A1) and were included in spectral analyses; 69 had C3:A2 available and were included in spindle analyses. PSG was recorded with the SOMNOscreen Plus system; manual scoring followed AASM criteria (Iber, 2007). Zmax recorded F7-Fpz (EEGL) and F8-Fpz (EEGR) at 256 Hz. All recordings were performed in participants’ own homes.

### 2.2 Signal Processing

For each 30-second epoch, log10-transformed bandpower was computed using Welch’s method with 4-second windows, for five frequency bands, defined following standard sleep EEG conventions (Iber, 2007): delta (0.5–4 Hz), theta (4–8 Hz), alpha (8–12 Hz), sigma (12–16 Hz), and beta (16–30 Hz) and 50% overlap, providing a frequency resolution of 0.25 Hz; sufficient to resolve the delta band lower boundary at 0.5 Hz while maintaining adequate spectral stability per epoch. The 4-second window was selected as the shortest window providing reliable delta power estimation, consistent with recommendations for sleep EEG spectral analysis at standard sampling rates (Iber et al., 2007; Vallat & Walker, 2021). A sensitivity analysis comparing 4-second and 8-second windows confirmed that window length had negligible effects on alpha, theta, sigma, and beta power estimates (mean difference <0.009 log_10_ units across bands). Delta power showed a systematic positive offset with the 4-second window relative to 8-second (mean difference=0.092 log_10_ units, SD=0.057), attributable to reduced low-frequency resolution and spectral leakage from sub-delta components. As both Zmax and PSG channels were processed with identical parameters, this offset cancels in all comparative analyses; absolute delta power values should not be compared to norms derived with different window lengths. Epoch stage labels were taken exclusively from PSG manual scoring, never from automated wearable staging consistent with the recommendation from our companion staging validation study. Features were extracted from the PSG frontal left channel (F3:A2), PSG central left channel (C3:A2), and both Zmax channels (F7-Fpz, F8-Fpz).

### 2.3 Per-Subject N2 Calibration

To correct for the systematic bandpower offset introduced by the active Fpz reference, a per-subject calibration offset was computed for each frequency band as the difference between the subject’s mean PSG frontal and Zmax bandpower during PSG-defined N2 epochs (Nunez & Srinivasan, 2006). This offset was then added to all Zmax bandpower values for that subject across all stages and bands. N2 was selected as the calibration reference stage because it provides the most epochs per subject (Carskadon & Dement, 2011; Stores, 2001), has the most stable spectral profile, and showed the highest pre-calibration Spearman correlations. Calibration was validated on held-out stages (Wake, N1, N3, REM) using Bland-Altman analysis (Chinoy et al., 2021; Martin Bland & Altman, 1986) and Spearman correlation.

### 2.4 Lateralisation Index Analysis

Lateralisation indices (LI) were computed as (Left − Right) / (Left + Right) per band per stage per subject for both Zmax (F7 vs F8) and PSG channels. Two PSG reference schemes were tested: bipolar-referenced (F3:A2 vs F4:A1) and common-referenced (F3 vs F4, unipolar), the latter matched to the shared-reference Zmax design. LI correspondence between Zmax and PSG was assessed by Spearman correlation.

### 2.5 Sleep Spindle Detection

Spindle detection was performed using YASA(Vallat & Walker, 2021) version 0.6.5 on three channels per subject: PSG C3:A2 (canonical AASM channel), PSG F3:A2 (frontal topographic match for Zmax), and Zmax F7-Fpz (EEGL). Detection parameters followed YASA defaults: sigma band 12–15 Hz, minimum duration 0.5 s, maximum duration 2.0 s, minimum inter-spindle distance 500 ms. Detection was restricted to PSG-defined N2 and N3 epochs. Spindle density was expressed as spindles per minute of N2+N3 time. Correspondence between channels was assessed using Pearson and Spearman correlation and Bland-Altman analysis.

## 3. Results

### 3.1 Systematic Bandpower Underestimation

Bland-Altman analysis of N2 bandpower revealed systematic underestimation of all frequency bands by Zmax relative to PSG frontal (F3:A2). Bias values were: delta −0.41 log units, theta −0.70, alpha −0.69, sigma −0.74, beta −0.44 (all negative, indicating Zmax < PSG). In linear terms, the sigma band bias of −0.74 log units corresponds to approximately 5.5× lower absolute power in Zmax relative to PSG frontal. The systematic direction of underestimation across all bands is consistent with the active Fpz reference introducing partial common-mode cancellation of cortical signals. Per-subject calibration offsets showed substantial inter-individual variability (sigma: SD=0.296), indicating that a single global correction is insufficient and per-subject calibration is required.

**Figure 1.**
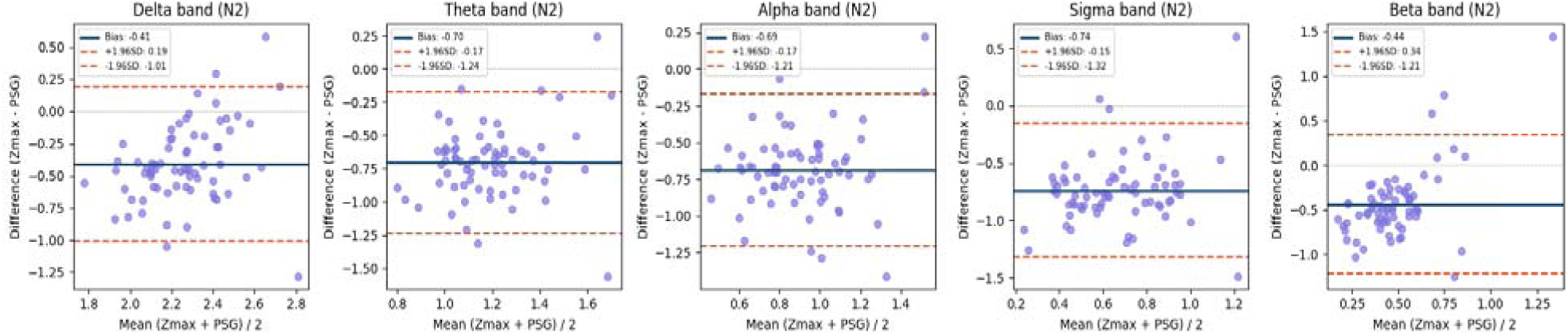
Systematic Zmax bandpower underestimation across all frequency bands before calibration.

Bland-Altman plots for Zmax versus PSG frontal left channel log bandpower during PSG-defined N2 epochs (N=71 subjects), for delta, theta, alpha, sigma, and beta bands. Each point represents one subject. Horizontal lines show mean bias (blue) and 95% limits of agreement (red dashed). All bands show systematic negative bias (Zmax < PSG), ranging from –0.41 log units for delta to –0.74 log units for sigma, consistent with partial signal cancellation introduced by the active Fpz reference electrode.

### 3.2 Pre-Calibration Spectral Correspondence

Before calibration, Zmax bandpower showed moderate but significant correspondence with PSG frontal bandpower at the subject level (N=71, Spearman correlations). N2 showed the strongest pre-calibration correlations: alpha (r=0.556, p<0.001), sigma (r=0.512, p<0.001), theta (r=0.430, p<0.001). N3 alpha was the strongest single pre-calibration correlation across all stage-band combinations (r=0.646, p<0.001). Beta was notably weaker: non-significant in N2 (r=0.055, p=0.65) and N3 (r=0.031, p=0.80). The left Zmax channel (F7-Fpz) consistently outperformed the right channel (F8-Fpz) across stage-band combinations.

**Figure 2.**
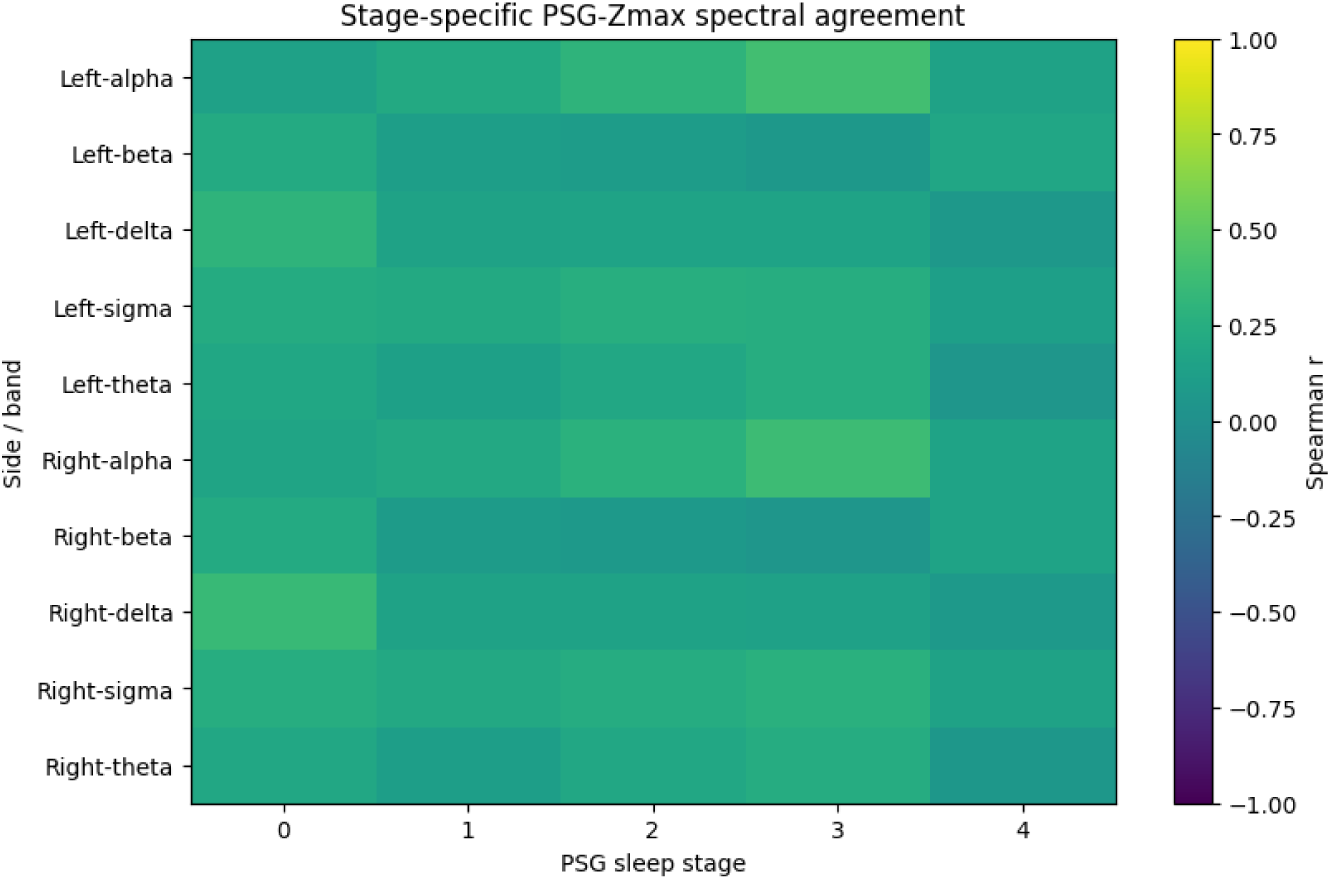
Stage-specific Zmax-to-PSG spectral agreement before calibration.

Heatmap of Spearman r between Zmax and PSG frontal log bandpower across five frequency bands and five sleep stages, for left (F7-Fpz) and right (F8-Fpz) Zmax channels (N=71). Colour scale ranges from –1 (dark blue) to +1 (yellow). All pre-calibration correlations are positive and moderate, with N2 showing the strongest correspondence. The left channel consistently shows higher agreement than the right channel across stage-band combinations, consistent with the left-channel advantage reported in the companion staging validation study.

### 3.3 Post-Calibration Spectral Correspondence

Per-subject N2-referenced calibration eliminated the systematic bias (post-calibration bias ≈0.000 for all bands, confirmed by Bland-Altman analysis). Spearman correlations in held-out stages improved substantially across all bands. N3 showed the largest improvements: alpha r=0.806 (pre: 0.646), sigma r=0.752 (pre: 0.487), theta r=0.734 (pre: 0.493), delta r=0.586 (pre: 0.278). REM alpha improved to r=0.755 (pre: 0.435) and REM sigma to r=0.667 (pre: 0.330). Beta, which was non-significant before calibration in N3 (r=0.031), reached r=0.546 after calibration. All post-calibration correlations in held-out stages were statistically significant (p<0.01).

**Figure 3.**
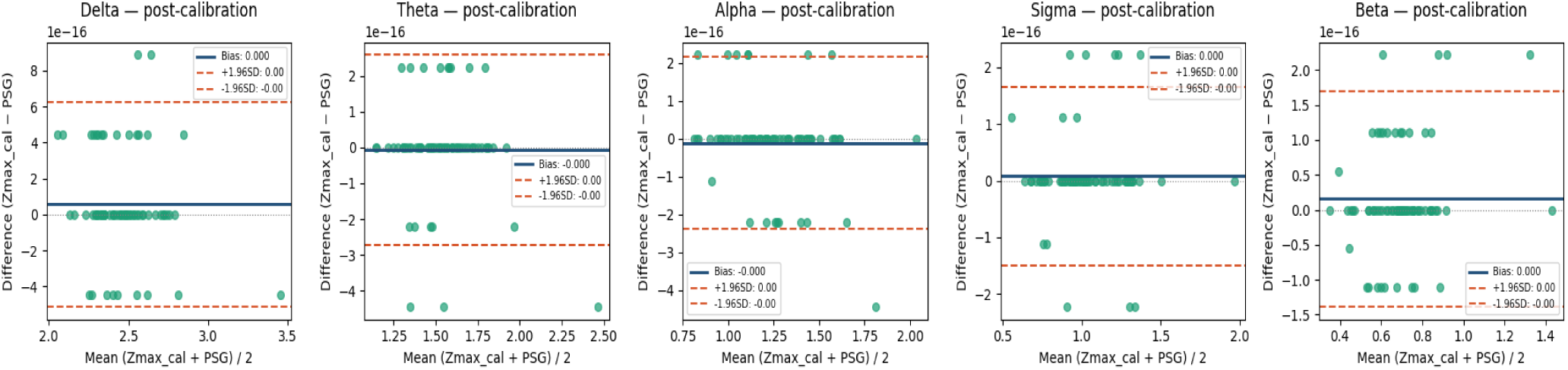
Elimination of systematic bias after per-subject N2 calibration.

Bland-Altman plots for calibrated Zmax versus PSG frontal log bandpower during PSG-defined N2 epochs (N=71 subjects). The per-subject N2 calibration offsets eliminate the systematic bias by construction (bias ≈ 0 for all bands, values on the order of 10^-16^ reflecting floating-point precision).

**Table 1.** Systematic Zmax bandpower underestimation (N2, left channel, N=71 subjects).

| Band | Bias (log10) | LoA lower | LoA upper | Interpretation |
| --- | --- | --- | --- | --- |
| Delta | -0.41 | -1.01 | +0.19 | Moderate attenuation |
| Theta | -0.70 | -1.24 | -0.17 | Strong attenuation |
| Alpha | -0.69 | -1.21 | -0.17 | Strong attenuation |
| Sigma | -0.74 | -1.32 | -0.15 | Strong attenuation;<br>~5.5× lower absolute<br>power |
| Beta | -0.44 | -1.21 | +0.34 | Moderate attenuation;<br>widest LoA |

**Table 2.** Pre– and post-calibration Spearman correlations: Zmax vs PSG frontal (left channel, subject level, N=71).

| Stage | Band | Pre r | Pre p | Post r | Post p | $\Delta r$ | N |
| --- | --- | --- | --- | --- | --- | --- | --- |
| N3 | alpha | 0.646 | <0.001 | 0.806 | <0.001 | +0.160 | 70 |
| N3 | sigma | 0.487 | <0.001 | 0.752 | <0.001 | +0.265 | 70 |
| N3 | theta | 0.493 | <0.001 | 0.734 | <0.001 | +0.241 | 70 |
| REM | alpha | 0.435 | <0.001 | 0.755 | <0.001 | +0.320 | 70 |
| REM | sigma | 0.330 | 0.005 | 0.667 | <0.001 | +0.337 | 70 |
| N2 | sigma | 0.512 | <0.001 | 1.000 | N/A | [calib]* | 71 |
| N3 | beta | 0.031 | 0.797 | 0.546 | <0.001 | +0.515 | 70 |

N2 post-calibration $r=1.000$ is by construction (calibration reference stage) and is excluded from held-out validation. $\Delta r = \text{post} - \text{pre calibration Spearman } r$ .
\*calibration reference stage: $r=1.000$ by construction, excluded from held-out validation

### 3.4 Robustness of N2 Calibration: N3 and REM Reference Stages Comparison

To verify that the N2-derived calibration offset generalises across stages, the calibration procedure was repeated using N3 and REM as alternative reference stages, with held-out performance evaluated on all remaining stages. N2 calibration produced the highest mean post-calibration Spearman correlation across all bands and validation stages (r□=0.601), outperforming N3 calibration (r□=0.479) and REM calibration (r□=0.489). Critically, N2 calibration produced zero incremental change in held-out Spearman r (Δr =0.000 for all band-stage combinations), indicating that the N2-derived offset accurately characterises the cross-stage reference bias without stage-specific distortion. N3 and REM calibrations introduced systematic over-or under-correction in held-out stages, reducing delta Spearman r by up to 0.676 points (REM-N2) and 0.441 points (N3-N2) respectively. The N2 calibration advantage was most pronounced for delta (N2: r=0.490 vs N3: 0.372, REM: 0.304) and sigma (N2: r=0.640 vs N3: 0.511, REM: 0.637), the two bands most relevant for sleep biomarker research. These results confirm that N2 is the optimal calibration anchor for Zmax spectral correction, attributable to its central position in the sleep spectral landscape between the delta-dominant N3 and mixed-frequency Wake/REM stages.

### 3.5 Within Subject Reliability, Inter-Individual Variability, and Demographic Independence of Spectral Offsets

Within-subject reliability of calibrated Zmax N2 bandpower was assessed using split-half correlation across N2 epochs within each recording night. Spearman-Brown corrected split-half reliability was excellent for all five frequency bands (delta: r_SB=0.995; theta: r_SB=0.996; alpha: r_SB=0.997; sigma: r_SB=0.998; beta: r_SB=0.998), indicating that mean bandpower estimates derived from any half of the available N2 epochs are highly predictive of estimates derived from the other half. Epoch-level within-subject coefficient of variation was low across all bands (delta: mean CV=7.0%, median=5.5%; sigma: mean CV=3.6%, median=2.4%; alpha: 3.9%; theta: 4.3%; beta: 3.8%), consistent with stable physiological signal rather than measurement noise.

One-way ICC(1,1) values computed from variance components were low across bands (delta: 0.126; theta: 0.172; alpha: 0.218; sigma: 0.257; beta: 0.311). This pattern reflects large between-subjects variance in absolute bandpower (BMS=28–51) relative to within-subjects variance (WMS=0.25–0.57) rather than poor measurement reliability. Between-subjects EEG spectral variance is biologically expected as individuals differ substantially in baseline cortical activity, and is the signal of interest for biomarker applications. The low ICC(1,1) therefore indicates that per-subject calibration is essential before cross-subject comparison, consistent with the calibration framework proposed here.

To assess whether inter-individual variability in spectral offsets is attributable to demographic factors, Spearman correlations between the pre-calibration PSG-Zmax offset and age and sex were computed across all bands and stages. No significant associations were found (all p>0.077; maximum R^2^=0.038 for N2 sigma). Age and sex together explained less than 4% of offset variance in any band, indicating that the inter-individual variability in spectral offsets reflects subject-specific hardware-interface properties; including electrode contact geometry, scalp impedance, and skull characteristics, rather than demographic factors accessible without dedicated measurement. Together, these results indicate that calibrated Zmax spectral features show excellent within-subject stability suitable for longitudinal monitoring, while requiring personalised per-subject calibration that cannot be approximated by demographic adjustment.

### 3.6 Lateralisation Analysis

Lateralisation indices (LI = left minus right channel bandpower) showed no significant correlation with PSG reference measures in any band or stage (N2 delta: r=0.080, 95% CI [-0.156, 0.308]; N2 sigma: r=0.050, 95% CI [-0.185, 0.280]; N3 delta: r=0.120, 95% CI [-0.122, 0.348]; REM alpha: r=0.090, 95% CI [-0.157, 0.327]). However, all confidence intervals extended into moderate effect territory (upper bounds r=0.28– 0.35), and post-hoc power analysis indicated only 36–39% power to detect a small-to-moderate effect of r=0.20 at the observed sample sizes (N=65–71), with N=194 required for 80% power at a=0.05. These findings should be interpreted as absence of evidence rather than evidence of absence: the present study was insufficiently powered to exclude clinically meaningful hemispheric asymmetry in Zmax frontal EEG.

**Figure 4.**
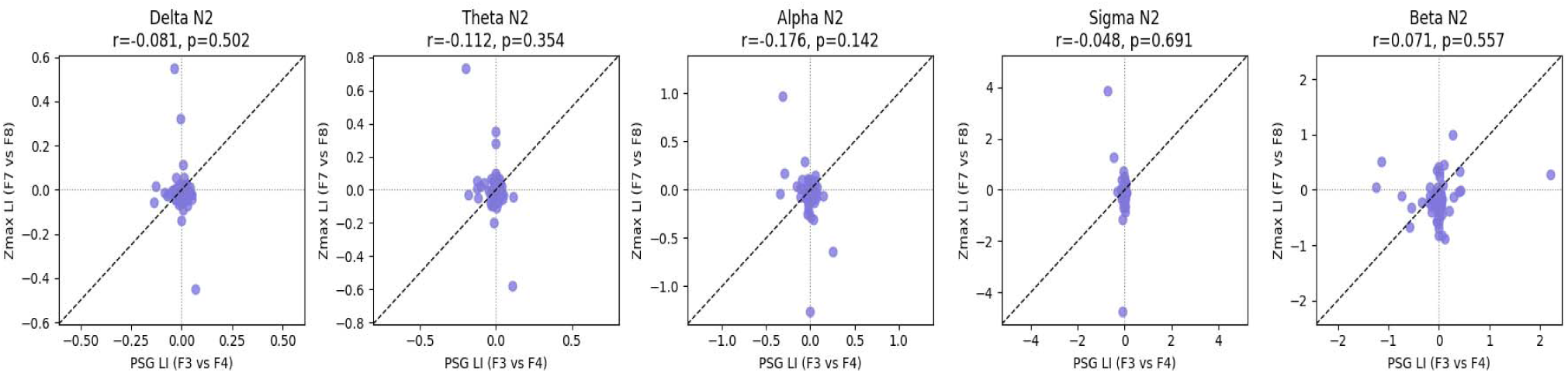
Null lateralisation index correspondence between Zmax and PSG frontal channels.

Scatter plots of Zmax lateralisation index (F7 minus F8, divided by sum) versus PSG lateralisation index (F3 minus F4, divided by sum) for five frequency bands during N2 (N=71 subjects). Spearman r and p-values shown above each panel. The identity line (dashed) is shown for reference. All correlations are near zero and non-significant (all p>0.14), with Zmax lateralisation indices clustered near zero regardless of PSG lateralisation magnitude, consistent with insufficient inter-electrode distance for hemispheric asymmetry detection.

### 3.7 Sleep Spindle Detection

Spindle detection (N=69 subjects with C3:A2 available) yielded mean spindle densities of 2.06±1.05 spindles/min for PSG C3, 1.62±0.98 for PSG F3, and 1.01±0.98 for Zmax F7. This three-tier attenuation pattern (PSG C3 > PSG F3 > Zmax F7) is mechanistically consistent with spindle topography: fast spindles are centroparietal-dominant and progressively attenuated at frontal and frontotemporal electrodes, with the active Fpz reference introducing additional sigma-band attenuation in Zmax.

As shown on Figure 5, PSG C3 vs PSG F3 showed high agreement (Pearson r=0.863, Spearman r=0.849), providing an internal reliability benchmark. PSG F3 vs Zmax F7 showed moderate-to-strong agreement (Pearson r=0.606, Spearman r=0.683, both p<0.001), indicating that the rank ordering of individual spindle density is substantially preserved in Zmax. PSG C3 vs Zmax F7 showed moderate agreement (Pearson r=0.494, Spearman r=0.551, both p<0.001). Bland-Altman analysis for PSG F3 vs Zmax F7 revealed a systematic bias of −0.62 spindles/min (LoA: −2.32 to +1.09), confirming the under-detection pattern. The PSG C3 vs Zmax F7 bias was larger (−1.05 spindles/min, LoA: −3.05 to +0.96), reflecting both topographic and reference attenuation.

**Figure 5.**
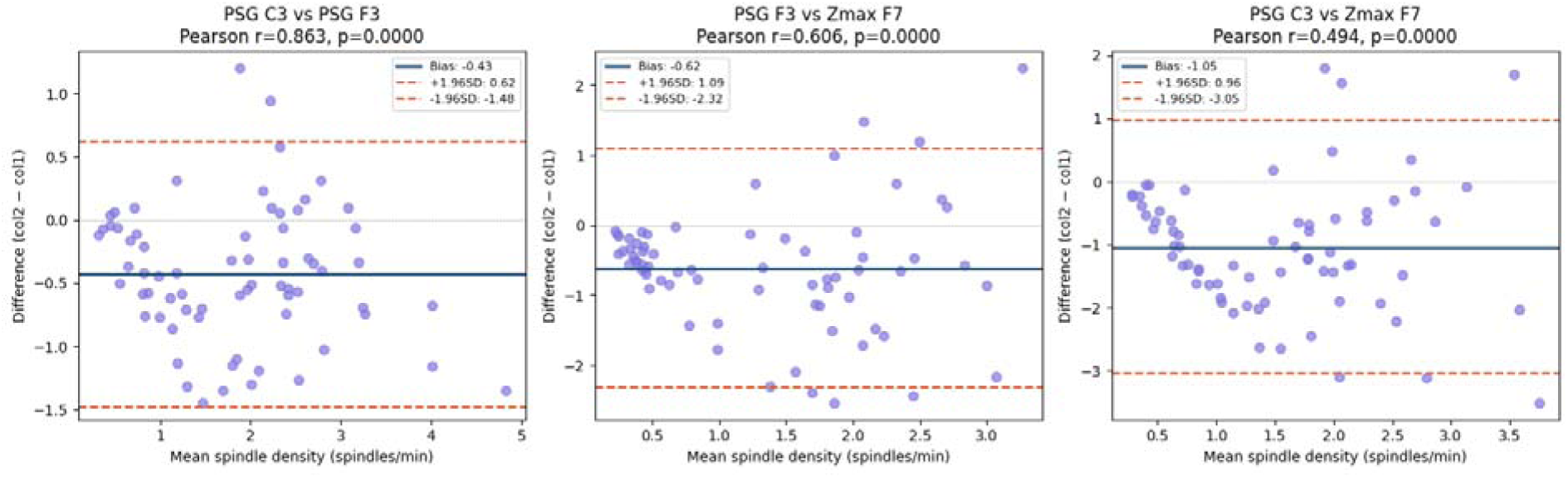
Bland-Altman analysis of sleep spindle density across three channel comparisons.

**Table 3.** Sleep spindle density: PSG C3, PSG F3, and Zmax F7 (N2+N3, YASA, N=69 subjects).

| Comparison | Pearson r (p) | Spearman r (p) | Bias<br>(spindles/min) | LoA (95%) |
| --- | --- | --- | --- | --- |
| PSG C3 vs<br>PSG F3 | 0.863 (<0.001) | 0.849 (<0.001) | $-0.43$ | $-1.48$ to $+0.62$ |
| PSG F3 vs<br>Zmax F7 | 0.606 (<0.001) | 0.683 (<0.001) | $-0.62$ | $-2.32$ to $+1.09$ |
| PSG C3 vs<br>Zmax F7 | 0.494 (<0.001) | 0.551 (<0.001) | $-1.05$ | $-3.05$ to $+0.96$ |

A parameter sensitivity analysis was conducted to determine whether spindle under-detection was algorithmic or hardware-driven (shown on Table 4). At F7-Fpz, mean N2 relative sigma power was 0.174 across subjects, falling below YASA’s default relative power pre-filter threshold of 0.20 which was calibrated on central PSG channels where sigma dominates. Reducing relative power to 0.05 recovered PSG F3-equivalent absolute density (mean Zmax=1.692 sp/min, bias=−0.067 vs PSG F3, Spearman r=0.685), confirming the under-detection is attributable to algorithmic pre-filtering rather than hardware amplitude limitations. Spearman r showed minimal sensitivity to threshold (range 0.669–0.685 across all tested parameters), indicating that rank-order correspondence is robust regardless of the absolute count threshold used.

**Table 4.** Parameter Sensitivity Analysis of Relative Sigma Power.

| Relative<br>Sigma<br>Power | Spindle<br>Frequency | Mean<br>Zmax | Bias vs F3 | Spearman r |
| --- | --- | --- | --- | --- |
| 0.05 | (12,15) | 1.692 | -0.067 | 0.685 |
| 0.08 | (12,15) | 1.574 | +0.051 | 0.669 |
| 0.10 | (12,15) | 1.488 | +0.136 | 0.671 |
| 0.12 | (12,15) | 1.395 | +0.230 | 0.676 |
| 0.15 | (12,15) | 1.254 | +0.371 | 0.680 |
| **0.20**<br>**(default)** | **(12,15)** | **1.009** | **+0.616** | **0.683** |
| 0.10 | (12,16) | 1.531 | +0.094 | 0.641 |
| 0.10 | (11,16) | 2.056 | -0.432 | 0.619 |

## 4. Discussion

### 4.1 The Active Fpz Reference as the Primary Source of Systematic Bias

The universal negative bias in Zmax bandpower (−0.41 to −0.74 log units) is mechanistically explained by the active Fpz reference. In standard EEG bipolar referencing, both F7 and F8 are measured relative to Fpz, which sits over prefrontal cortex and therefore carries its own EEG activity. Subtracting an active reference attenuates the apparent power of the active electrode because part of the target signal is cancelled by the common-mode reference signal. This effect is systematic and consistent across all frequency bands, explaining the near-uniform bias pattern in Table 1. PSG referencing to the mastoid (A1, A2) avoids this problem because the mastoid is electrically remote from cortical generators. The magnitude of the bias (≈0.6 log units for theta, alpha, sigma) is consistent with theoretical predictions for active-reference subtraction when both electrode and reference sites share significant spectral energy.

The inter-individual variability in calibration offsets (SD ≈0.27–0.39 log units across bands) confirms that a single global correction is insufficient. Per-subject calibration is necessary because the degree of reference contamination depends on individual factors including skull thickness, skin-electrode impedance, and the spatial distribution of each subject’s cortical generators. The N2-based calibration framework presented here provides a principled, reproducible method for per-subject correction that requires only a single overnight recording. N2 is the optimal calibration reference because it is the most abundant stage, has the most stable spectral signature, and shows the highest pre-calibration correlations as confirmed by the calibration robustness analysis in Section 3.4.

### 4.2 Post-Calibration Validity for Sleep Biomarker Research

After calibration, Zmax spectral features showed strong correspondence with PSG frontal features in held-out stages, particularly for alpha (N3 r=0.806, REM r=0.755) and sigma (N3 r=0.752, REM r=0.667). These are the two bands most relevant for sleep biomarker research: sigma captures sleep spindle activity related to memory consolidation and psychiatric phenotypes, and alpha during NREM reflects arousal intrusions associated with sleep quality and psychiatric vulnerability. The strength of these correlations (r>0.75) indicates that calibrated Zmax features are valid proxies for PSG frontal spectral features at the inter-individual level and the subjects who have more sigma or alpha power according to PSG also have more according to Zmax after calibration.

Delta during N3 showed a more moderate post-calibration correlation (r=0.586). This is somewhat surprising given that slow-wave activity is frontally dominant and should be well-captured by F7-Fpz. The modest improvement despite calibration likely reflects that N3 delta is a non-stationary signal with large epoch-to-epoch variability, making mean-level subject profiles less stable than for sigma or alpha. Studies using N3 delta as a primary biomarker should treat the Zmax estimates with more caution than sigma or alpha.

Beta reached r=0.546 in N3 and r=0.703 in REM after calibration, despite being near-zero before calibration. This improvement is partially attributable to the calibration anchoring the absolute power scale, which improves rank-order stability. However the large pre-calibration LoA for beta (Table 1) and its known susceptibility to EMG contamination and high-frequency noise at dry electrodes means beta should be used cautiously and only after explicit artefact screening.

As reported in Section 3.5, the dissociation between excellent within-subject reliability (r_SB>0.99) and low ICC(1,1) (0.13–0.31) has a direct practical implication for study design: Zmax is well-suited to within-subject longitudinal tracking but requires individual calibration for cross-subject comparisons. On the other hand, the inter-individual variability in post-calibration residual offsets (N3 delta SD=0.399 log units, range – 1.12 to +2.02) substantially exceeded the mean offset (0.082 log units), indicating that subject-specific hardware-interface factors dominate over systematic device-level bias. Age and sex explained less than 4% of this variance, consistent with the hypothesis that electrode-skin contact geometry; which varies with individual head shape, skin moisture, and hair distribution, is the primary source of inter-subject spectral variability. Future studies should consider including scalp impedance measurements as a covariate in spectral analyses to account for this variability.

The superiority of N2 calibration over N3 and REM alternatives reflects the spectral position of N2 as the stage whose reference offset most closely approximates the cross-stage average. N3 calibration introduces positive delta over-correction (because N3 delta power is abnormally high, inflating the estimated offset) while REM calibration introduces negative delta under-correction. This stage-specificity of reference offsets has a practical implication: calibration stages should be selected based on their spectral centrality, not solely on epoch abundance, and N2 satisfies both criteria for most healthy adult recordings.

### 4.3 Spindle Density as a Validated Wearable Biomarker

The PSG F3 vs Zmax spindle density correlation (r=0.606 Pearson, r=0.683 Spearman) is the most practically important finding for downstream research. It establishes that Zmax captures the inter-individual variation in frontal spindle density well enough to be a useful population-level biomarker, even though absolute counts require bias correction. The moderate Pearson but higher Spearman correlation indicates a non-linear or outlier-influenced relationship in absolute density units; the rank ordering is better preserved than the linear scaling which is consistent with the fan-shaped Bland-Altman pattern suggesting proportional rather than fixed under-detection at higher densities.

The three-tier density gradient (PSG C3: 2.06, PSG F3: 1.62, Zmax F7: 1.01 spindles/min) provides a useful reference for future Zmax studies. Researchers using YASA default thresholds on Zmax data should expect approximately 50% of the spindle counts they would observe on a PSG central channel. The PSG C3 vs PSG F3 benchmark (r=0.863) establishes that even within PSG, frontal and central spindle counts differ substantially where the Zmax frontal result (r=0.606 vs PSG frontal) is therefore better understood relative to this within-PSG frontal correlation than relative to the C3 gold standard.

The approximately 50% spindle under-detection by Zmax relative to PSG central channels is consistent with the lower amplitude of frontal spindle expression (Mölle et al., 2002) and represents a systematic bias that can be corrected by a multiplicative scaling factor estimated from the calibration session. Researchers using Zmax spindle density as a biomarker should apply this correction before comparing values to PSG-derived norms.

Spindle detection parameter sensitivity analysis demonstrated that YASA’s default rel_pow threshold (0.20) systematically suppresses frontal EEG spindle detection because frontal N2 relative sigma power (mean=0.174) falls below this PSG-central-calibrated threshold. Lowering rel_pow to 0.05 recovers PSG F3-equivalent absolute counts with near-zero bias, providing a validated parameter recommendation for Zmax spindle studies.

### 4.4 Lateralisation: An Underpowered Null Result with Design Implications

The absence of significant lateralisation index correspondence across all bands and stages must be interpreted cautiously in light of the power analysis reported in Section 3.6. With 36–39% power to detect r=0.20 and confidence intervals extending to r=0.28–0.35, the present data cannot exclude clinically meaningful hemispheric asymmetry; the null result reflects insufficient statistical power as much as genuine absence of lateralisation signal. A bigger sample would be required to test this question definitively. Researchers should not conclude from the present findings that Zmax is incapable of detecting hemispheric asymmetry; rather, this question remains open pending an adequately powered study.

That said, there are principled geometric reasons to expect limited lateralisation sensitivity. The Zmax F7-Fpz and F8-Fpz derivations are separated by approximately 6 cm of forehead surface; substantially less than the 10–15 cm inter-electrode distances used in validated frontal alpha asymmetry paradigms. At this inter-electrode distance, the differential signal between left and right channels is likely attenuated relative to the shared frontal field, reducing sensitivity to genuine hemispheric differences. This geometric constraint is a property of the current hardware design rather than a fundamental limitation of wearable EEG. Adding mastoid reference electrodes, as implemented in the Dreem headband’s F7-O1 and F8-O2 derivations or including central channels would substantially increase effective inter-electrode distance and likely improve lateralisation sensitivity. Future Zmax validation studies with larger samples and concurrent multi-electrode reference should explicitly test whether frontal alpha asymmetry or lateralised spindle density can be reliably captured under optimal conditions.

For researchers currently using Zmax, the practical recommendation is to treat the left and right channels as independent spectral measurements rather than an asymmetry pair. Each channel provides a separate estimate of frontotemporal EEG power, and bilateral recording therefore doubles the available spectral features relative to a single-channel device. Left-channel features showed marginally superior staging agreement with PSG in the present cohort (Section 3.6), motivating the recommendation to use F7-Fpz as the primary channel for algorithms requiring a single input. Asymmetry-based phenotyping, including frontal alpha asymmetry and lateralised spindle analyses should be treated as exploratory in Zmax data until adequately powered validation is available.

### 4.5 Validated Feature Set for Downstream Analyses

Based on the totality of findings, the following feature set is validated for use in downstream analyses of Wearanize+ Zmax data: (1) calibrated log bandpower for delta, theta, alpha, and sigma across Wake, N1, N3, and REM, using PSG-defined stage epochs and per-subject N2-referenced calibration; (2) spindle density (spindles/min of N2+N3) with acknowledgement of systematic under-detection relative to PSG frontal and central channels; (3) left and right channel features treated independently, without lateralisation index computation. Beta power may be included with explicit caveats regarding its pre-calibration instability and susceptibility to artefact.

### 4.6 Limitations

Several limitations apply. First, calibration was performed and evaluated within the same single-night recording, precluding assessment of calibration stability across nights. Published evidence from repeated PSG studies suggests that EEG spectral profiles show moderate night-to-night reliability (ICC=0.60–0.85 for delta and sigma power)(Brunner et al., 1993), which would support calibration stability if the offset reflects stable hardware-subject interface properties. However, if electrode contact quality varies night-to-night, recalibration may be required for each recording session. Future multi-night studies should explicitly test whether a single calibration session can be reused across subsequent Zmax recordings without performance degradation. Second, The N2-based calibration approach assumes sufficient N2 epochs for stable offset estimation. In clinical populations with severely disrupted sleep architecture, including insomnia disorder (reduced N2 continuity), schizophrenia (reduced sleep spindles), and advanced age (N2 fragmentation) and the available N2 epoch count may be insufficient for reliable calibration. Alternative calibration strategies warrant investigation: cumulative NREM calibration (pooling N1, N2, and N3 epochs) would maximise available data but introduce stage-mixing; wake calibration (eyes-closed resting EEG before sleep onset) would avoid stage-dependency entirely but assumes spectral stability between wakefulness and sleep. These alternatives should be evaluated in clinical cohort studies before the Zmax calibration framework is applied outside healthy community samples. Third, the generalisability of the N2 calibration framework to older adults and clinical populations with altered sleep architecture requires specific consideration. In healthy older adults, N2 sleep is characterised by reduced spindle amplitude and density, flattening of the sigma peak, and increased low-frequency intrusions, which may systematically alter the spectral centroid of the N2 reference epoch and introduce age-dependent miscalibration. In patients with obstructive sleep apnea, N2 epochs are interrupted by arousal-driven power transients that inflate broadband power estimates and would corrupt offset estimation if not explicitly excluded. In insomnia disorder, where N2 is often fragmented and subjectively misperceived as wakefulness, staging errors would propagate directly into the calibration epoch pool. For these populations, alternative calibration strategies may be necessary: eyes-closed pre-sleep resting EEG would circumvent stage-dependency entirely, while cumulative NREM pooling would increase epoch count at the cost of stage-mixing. Researchers extending the present framework to clinical cohorts should validate calibration performance against concurrent PSG in each target population before applying N2-referenced corrections developed in healthy young adults. Fourth, spindle detection used YASA default parameters not optimised for frontal EEG; algorithm tuning specifically for F7-Fpz derivations may improve detection accuracy. Fifth, three subjects with non-standard PSG montages were excluded, limiting generalisability to variants of the Wearanize+ collection protocol.

## 5. Conclusions

This study provides a comprehensive spectral validation framework for the Zmax EEG headband using the Wearanize+ home-recording dataset. The key contributions are: (1) characterisation and mechanistic explanation of systematic bandpower underestimation (−0.41 to −0.74 log units) across all frequency bands; (2) a per-subject N2-referenced calibration protocol that eliminates this bias and substantially improves held-out stage spectral correspondence (N3 alpha r=0.806, N3 sigma r=0.752); (3) validation of spindle density as a wearable biomarker (PSG F3 vs Zmax r=0.606), with quantification of systematic under-detection; and (4) A qualified null result for lateralisation analysis, with confidence intervals extending to r=0.28–0.35 indicating the study was insufficiently powered to exclude clinically meaningful hemispheric asymmetry. These findings establish the methodological prerequisites for wearable-based sleep biomarker research using Wearanize+, including the multi-omics integration studies planned as direct follow-ons. One technical caveat warrants explicit statement: absolute delta power values reported throughout this study were computed using 4-second FFT windows. As described in Section 2.2, this introduces a systematic positive offset relative to norms derived from 8-second windows. Comparative and cross-study analyses should account for this discrepancy; the present absolute delta values should not be cited as normative references for studies using longer window lengths. Until cross-night calibration stability is empirically established, the framework should be applied within-session which is the N2-referenced offset estimated from a given recording night and should be used exclusively to calibrate spectral features from that same night rather than carried forward as a portable correction profile. Studies requiring longitudinal tracking of spectral biomarkers across multiple nights should plan for per-session recalibration as a default, and should treat between-session spectral differences as potentially confounded by night-to-night variation in electrode-skin contact geometry until multi-night stability data become available.

## Data Availability Statement

The Wearanize+ PlugNPlay dataset used in this study is publicly available via the Radboud Data Repository under a signed Data Use Agreement permitting non-commercial academic research use. The dataset can be accessed at https://doi.org/10.1093/sleepadvances/zpaf094 (Sikder et al., 2026). Analysis code used to produce the results reported in this manuscript is available on request from the corresponding author.

## IRB Statement

This study used a publicly available secondary dataset (Wearanize+ PlugNPlay, Radboud Data Repository). All data collection, participant recruitment, and ethical oversight were conducted by the original dataset creators. The original study was approved by the relevant institutional ethics committee and all participants provided written informed consent prior to data collection, as described in Sikder et al. (2026). Secondary analysis of this anonymised publicly available dataset under a signed Data Use Agreement did not require additional ethical approval from the authors’ institutional review board.

